# The built environment is more predictive of cardiometabolic health than other aspects of lifestyle in two rapidly transitioning Indigenous populations

**DOI:** 10.1101/2024.08.26.24312234

**Authors:** Marina M. Watowich, Audrey M. Arner, Selina Wang, Echwa John, John C. Kahumbu, Patricia Kinyua, Anjelina Lopurudoi, Francis Lotukoi, Charles M. Mwai, Benjamin Muhoya, Boniface Mukoma, Kar Lye Tam, Tan Bee Ting A/P Tan Boon Huat, Michael Gurven, Yvonne A L Lim, Dino Martins, Sospeter Njeru, Ng Kee Seong, Vivek V. Venkataraman, Ian J. Wallace, Julien F. Ayroles, Thomas S. Kraft, Amanda J. Lea

## Abstract

**Background:** Many subsistence-level and Indigenous societies around the world are rapidly experiencing urbanization, nutrition transition, and integration into market-economies, resulting in marked increases in cardiometabolic diseases. Determining the most potent and generalized drivers of changing health is essential for identifying vulnerable communities and creating effective policies to combat increased chronic disease risk across socio-environmental contexts. However, comparative tests of how different lifestyle features affect the health of populations undergoing lifestyle transitions remain rare, and require comparable, integrated anthropological and health data collected in diverse contexts.

**Methods:** We developed nine scales to quantify different facets of lifestyle (e.g., urban infrastructure, market-integration, acculturation) in two Indigenous, transitioning subsistence populations currently undergoing rapid change in very different ecological and sociopolitical contexts: Turkana pastoralists of northwest Kenya (n = 3,692) and Orang Asli mixed subsistence groups of Peninsular Malaysia (n = 688). We tested the extent to which these lifestyle scales predicted 16 measures of cardiometabolic health and compared the generalizability of each scale across the two populations. We used factor analysis to decompose comprehensive lifestyle data into salient axes without supervision, sensitivity analyses to understand which components of the multidimensional scales were most important, and sex-stratified analyses to understand how facets of lifestyle variation differentially impacted cardiometabolic health among males and females.

**Findings:** Cardiometabolic health was best predicted by measures that quantified urban infrastructure and market-derived material wealth compared to metrics encompassing diet, mobility, or acculturation, and these results were highly consistent across both populations and sexes. Factor analysis results were also highly consistent between the Turkana and Orang Asli and revealed that lifestyle variation decomposes into two distinct axes–the built environment and diet–which change at different paces and have different relationships with health.

**Interpretation:** Our analysis of comparable data from Indigenous peoples in East Africa and Southeast Asia revealed a surprising amount of generalizability: in both contexts, measures of local infrastructure and built environment are consistently more predictive of cardiometabolic health than other facets of lifestyle that are seemingly more proximate to health, such as diet. We hypothesize that this is because the built environment impacts unmeasured proximate drivers like physical activity, increased stress, and broader access to market goods, and serves as a proxy for the duration of time that communities have been market-integrated.

## Introduction

In low- and middle-income countries (LMIC) around the world, many populations are transitioning from small-scale, subsistence-level practices to more industrialized, urban lifestyles. These lifestyle changes are multifaceted and include integration into cash-dependent market-economies (i.e., market-integration), acculturation and assimilation with neighboring cultures, and increased access to built infrastructure (i.e., urbanization), resulting in drastic dietary, nutritional, economic, infrastructural, and social changes^1–3^. Importantly, many rapidly transitioning populations are Indigenous and/or marginalized, with transitions precipitated by structural and systemic processes such as colonialism, loss of land rights, and environmental degradation^3–6^. Urbanization and acculturation of subsistence-level and Indigenous groups has been justified and promoted because such changes often accompany increased access to modern healthcare and reductions in certain types of infectious disease, extreme poverty, and food insecurity, yet lifestyle change has simultaneously created an epidemic of cardiometabolic diseases in these populations around the world^3,6–11^. Importantly, the details and drivers of these epidemics often differ: for example, political or economic pressure, political stability, legal security to Indigenous lands, and accessibility and desirability of Indigenous lands can influence the speed of their development and degradation^12^. Further, dietary shifts may depend on government policies (e.g., international trade involvement), market accessibility, extent of individual-level income change, and social pressures (e.g., alcohol acceptance)^11,13^.

As expected from broader epidemiological patterns, studies within individual countries or Indigenous populations exhibiting a gradient of subsistence-level-to-urban lifestyles point toward subsistence living being protective against cardiometabolic diseases^14–20^. Poor diet, exposure to pollution, changes in sleep patterns, built infrastructure, market goods, and occupations that reduce daily activity levels in more urban environments are expected to drive these negative health effects^21–27^. Further, this body of work has generally shown greater effects of urban exposure on health outcomes in females than males^28^. Although focused studies of individual countries or Indigenous populations can be illuminating, we lack a generalized framework for quantifying lifestyle transitions and their effects on cardiometabolic health across populations. Building such a framework requires comparable, individual-level data across multiple transitioning populations.

Operationalizing distinct facets of the lifestyle transition is necessary to determine how these different facets relate and which are most predictive of health. For example, McDade and Adair (2001) decomposed the urban environment into community-level versus individual-level factors, and later studies have investigated the influence of these, and other, lifestyle factors (e.g., material wealth, diet, market access) on specific aspects of cardiometabolic health^20,29,30^. However, previous population-specific studies have mostly focused on determining how just one or a few aspects of lifestyle drive a limited number of cardiometabolic health outcomes (e.g., diet^31^, physical activity^32^, subsistence/wage labor reliance^33^, material wealth vs. market access^20^, diet vs. material wealth^29^, socioeconomic status vs. acculturation^34^), and therefore are limited in their ability to establish a general framework for assessing which aspects of lifestyle variation most strongly affect cardiometabolic health in a comparative context.

Here, we investigate how diverse facets of urbanization (i.e., growth of cities and accompanying infrastructure), industrialization (i.e., transition to manufacturing economy), acculturation, and market-integration affect cardiometabolic health by bringing together two datasets from remote-living, subsistence-level Indigenous communities living in dramatically different ecological, economic, and sociopolitical contexts: the Turkana of northwest Kenya and Orang Asli of Peninsular Malaysia. East Africa and Southeast Asia remain globally understudied, and within their respective regions, the Turkana and Orang Asli live in remote areas further underrepresented in global and public health studies and initiatives^35^. We chose to work with these populations because they are experiencing rapid lifestyle transitions, and consequently, we were able to work with individuals of the same or similar genetic background spanning extreme gradients from traditional, subsistence lifestyles to fully industrialized and market-integrated lifestyles^18,36^. We use integrated and coordinated data collection to investigate three questions. First, we compare how facets of the lifestyle transition are occurring in Turkana and Orang Asli using nine scales–three previously developed and six new–to capture a priori domains of lifestyle variation (e.g., diet, built environment, material wealth)^18,37,38^. Second, we investigate the extent to which these domains are associated with 16 cardiometabolic health phenotypes, compare patterns between populations, use sensitivity analyses to identify the most important individual components, and piecewise analyses to further characterize the effects of lifestyle transition. Third, we investigate whether these findings differ between females and males in both populations.

#### Research in context

##### Evidence before this study

We searched PubMed for studies published from database inception to June 24, 2024, that linked urbanicity to cardiometabolic health outcomes in subsistence-level populations using the search terms “(industrialization OR urbanicity) AND (Indigenous OR subsistence) AND cardiometabolic)” and found 32 results. No studies assessed the generalizability of findings across multiple countries or environmental contexts. Rarely is there comparable data to disentangle whether different features of lifestyle variation (e.g., market-integration, acculturation, infrastructure) affect transitioning populations in similar ways because this relies on comparable, integrated anthropological and health data across multiple subsistence-level, Indigenous societies.

##### Added value of this study

Determining the most potent and generalizable global drivers affecting health following urbanization and industrialization is essential for identifying vulnerable communities and creating effective policy, and to understand how context-specific these policies must be. This study integrates comparable data from two large-scale population studies of Indigenous societies experiencing rapid lifestyle change in very distinct environmental and sociopolitical contexts: the Turkana of Kenya and Orang Asli of Peninsular Malaysia. These populations are not only understudied but reside in regions underrepresented in the global and public health literature. Our analyses show that lifestyle change is occurring in different ways between the two populations, but urban infrastructure consistently better predicts cardiometabolic health than seemingly more-proximate features such as diet in both populations.

##### Implications of all the available evidence

Despite lifestyle transitions manifesting at different rates and in different ways across LMIC, we find that the same lifestyle features–local infrastructure and built environment–most strongly predict cardiometabolic health in two distinct Indigenous populations. In addition to the built environment, dietary changes comprise another, distinct axis of the lifestyle transition, suggesting that these features can be independently targeted for intervention. Further, lifestyle impacts cardiometabolic health similarly in both females and males, but females are generally more affected by urbanization, suggesting that interventions to mitigate the effect of urbanization on adiposity and related outcomes should provide greater support to females.

## Methods

### Turkana

The Turkana are traditionally a subsistence-level, nomadic pastoralist population from the remote Turkana Basin in northwest Kenya. Traditionally, approximately 80% of the Turkana diet has been derived from subsistence-level animal products including milk, meat, and blood^18,39^. Ongoing infrastructure construction and rapid economic development of Kenya has resulted in the growth of several urban centers in and near traditional Turkana lands and the expansion of small-scale markets. As a result, most Turkana no longer exclusively practice traditional pastoralism, instead relying on trade and increasingly participating in the market economy^18,40^. In addition to socioeconomic changes happening within the Turkana region, many Turkana have moved to highly urbanized areas in central Kenya in the last several decades, and now participate fully in the market-economy^18,40^. Compared to subsistence-level and remote-living individuals, Turkana living in urban areas in central Kenya have higher body fat, cholesterol, and blood pressure; however, it remains unknown how cardiometabolic phenotypes are associated with different specific facets of the lifestyle gradient^18,41^.

### Orang Asli

The term “Orang Asli” is an ethnonym referring broadly to the Indigenous peoples of Peninsular Malaysia (represented by at least 19 culturally distinct ethnolinguistic groups) with varied subsistence histories consisting of foraging, swidden agriculture, trade of rainforest products, or a combination thereof^42^. Orang Asli are multiethnic and typically categorized into three broad groups (the Negrito [Semang], Senoi, and Aboriginal Malay) who are genetically distinct but generally more genetically similar to each other than to ethnic Malays or other Asian populations^43^. Prior to the 1950’s, most Orang Asli lived in relatively small groups and relied primarily on subsistence economies. With rapid economic development in Malaysia, especially the expansion of rubber and oil palm agriculture, coupled with government programs aimed at integrating Orang Asli into the national economy, Orang Asli have been exposed to differing levels of lifestyle change^9,44^. Today, this manifests as a dramatic lifestyle gradient within a relatively small country: some Orang Asli villages are surrounded by urban or peri-urban development while others consist of small camps of highly mobile individuals occupying interior rainforest regions with a continued heavy reliance on subsistence foraging and farming^36^.

Sub-groups of the Orang Asli have been reported to have different levels of obesity and hypertension, but it remains unclear the extent to which identifiable aspects of lifestyle transitions directly influence these rates^45,46^.

### Cardiometabolic health biomarkers

The data we use herein comes from two ongoing projects, the Turkana Health and Genomics Project (THGP) and the Orang Asli Health and Lifeways Project (OA HeLP), previously described in Lea et al. (2020) and Wallace et al. (2022), respectively. As part of both projects, consenting adults (≥18 years) participated in structured survey data collection and the measurement of 14 cardiometabolic phenotypes, including waist circumference, body fat percentage, weight, BMI, waist-to-hip ratio, total cholesterol, high-density and low-density lipoprotein (HDL, LDL) cholesterol, non-HDL cholesterol, LDL-to-HDL ratio, triglycerides (mg/dL), glucose level (mg/dL), and systolic and diastolic blood pressure (mm Hg). We also characterized whether the individual fit criteria for being hypertensive (systolic blood pressure > 135 and diastolic blood pressure > 85) or overweight or obese (BMI ≥ 25) (Figure 1A).

**Figure 1:**
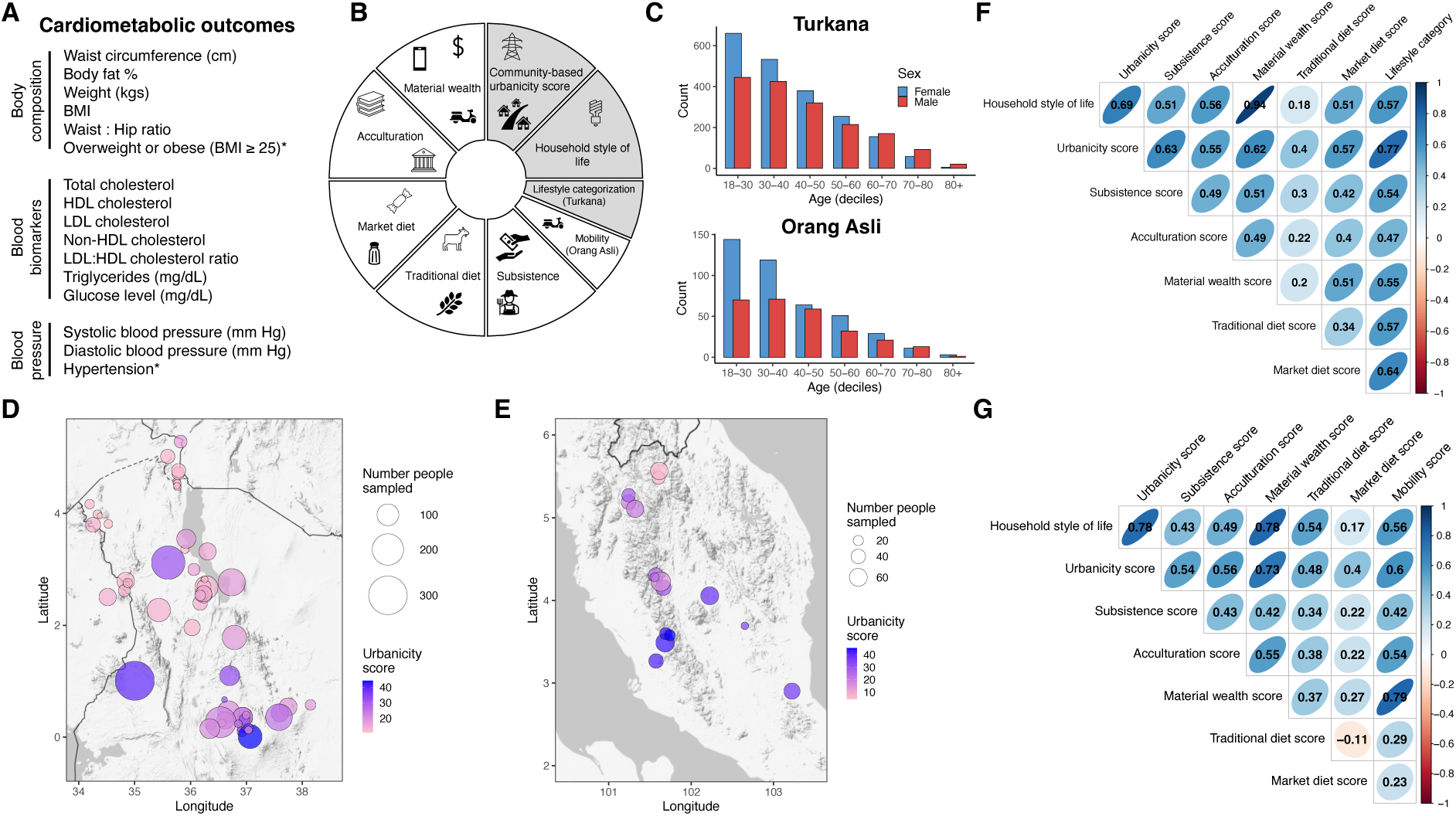
Study design overview. (A) The sixteen cardiometabolic traits we measured in both Turkana and Orang Asli. (B) Schematic of the nine scales used to quantify facets of urbanicity. Scales adapted from prior studies are colored gray. The two scales used for only one population are denoted. (C) Age and sex distributions of all individuals in the study. Maps showing the location-based urbanicity score for each sampling location in (D) Kenya and (E) Malaysia, with points scaled to the number of individuals sampled in each location. Correlations between all lifestyle variation scales in the (F) Turkana and (G) Orang Asli.

Development of scales to quantify particular facets of lifestyle variation We quantified lifestyle variation in both populations using nine scales which captured different facets of urbanicity, market-integration, dietary transitions, and acculturation (hereafter “lifestyle scales” or “scales”). Seven scales were replicated between the two populations and one was unique to each population (Figure 1B; Table S1). For both Turkana and Orang Asli, we adapted the previously published household style of life scale [H-SOL] from Gildner et al. (2020) and the location-based lifestyle scale from Novak et al. (2012)^37,38^. H-SOL measures an individual’s household size, building materials, and infrastructure (e.g., running water and electricity source). The location-based urbancity scale (hereafter “urbancity scale” or “urbanicity score”) captures population density and aggregate access to infrastructure (e.g., electricity source, sewage), material wealth in urban contexts, subsistence strategies, and education per location (Tables 1, S1). For Turkana, we also adopted a numeric version of the lifestyle categorization from Lea et al. (2020), which characterizes Turkana-specific subsistence strategies.

We then developed five scales that we a priori expected to capture different key facets of the lifestyle transition, including (i) material wealth, (ii) subsistence, which measures the extent to which individuals participate in wage labor vs. pastoralism, subsistence agriculture, hunting, fishing, or gathering, (iii) acculturation to the majority culture (e.g., speaking the majority language or practicing the religion of the predominant culture), and the extent to which individuals consume a (iv) traditional or (v) market-derived diet. Additionally, for Orang Asli only, we quantified individuals’ mobility and access to urban areas, as we expect that vehicle access in Malaysia is more indicative of market access than in Kenya given the size of the country (i.e., in Kenya, major urban centers are a multi-day drive from traditional Turkana lands). We note that the traditional diet scale is based on questions that quantify the extent of consumption of traditional foods but is coded such that higher values indicate less consumption of traditional foods. All lifestyle scales are coded such that higher values indicate higher extent of urbanicity, acculturation, or market-integration. All scales quantify lifestyle at the level of the individual except for the location-based urbanicity scale (which would be the same for all members of the same community).

To be clear, while many of the lifestyle scales we generated have the same range (0-1), we do not expect that the same value across different scales represents the same point in the lifestyle transition (Figure S1). Further, we did not design these scales to represent the lifestyle transition in absolute terms where the lowest number represents “fully” traditional, subsistence-level lifestyles and the highest number represents “fully” acculturated, urban, or market-integrated lifestyles. Instead, these scales were designed to capture relative variation in lifestyle features and be broadly applicable to multiple transitioning populations (Figure S1). After generating the lifestyle scales for each population, we compared the correlations between all scales for each population. Summary of age, sex, and lifestyle are available in Tables S2 and S3. Items included in each scale are listed in Table 1, with further details in Table S1. Code to generate each scale is available on our GitHub (https://github.com/mwatowich/Multi-population_lifestyle_scales).

**Table 1.** The nine lifestyle variation scales, the facets of lifestyle that each scale measured, and specific items included in each scale.

| Scale | Domain of lifestyle targeted | Items included |
| --- | --- | --- |
| Location-based urbanicity score (adapted from Novak et al. 2012) | Population density and aggregate infrastructure and material wealth (as defined in 'Western' contexts) per location | • Population density<br>• Proportion of houses with access to electricity, sewage<br>• Proportion of population engaged in subsistence practices<br>• Proportion of households with mobile phones, televisions<br>• Infrastructure available to access village (Orang Asli only)<br>• Proportion of individuals >40 years with some education, proportion of individuals <40 years with some education |
| Household style of life (H-SOL) (adapted from Gildner et al. 2020) | Household building materials and infrastructure | • Household size<br>• Household building materials<br>• Electricity, sewage, or tap/treated water infrastructure |
| Material wealth | Extent of wealth as defined in 'Western' or industrialized contexts | • Household size<br>• Household building materials<br>• Electricity, sewage, or tap/treated water infrastructure<br>• Ownership or access to mobile phone or TV |
| Acculturation | Extent of integration into majority culture of the country | • Highest education level<br>• Languages spoken<br>• Religion (Orang Asli only) |
| Market-derived diet | Extent of market-derived foods incorporated in diet | • Sugar, salt, and cooking oil consumption |
| Traditional diet | Extent (or lack thereof) of traditional foods consumed<br>*this scale is coded such that higher values indicate less consumption of traditional foods | • Turkana: meat, milk, blood consumption<br>• Orang Asli: rice, manioc, wild meat, and wild fish consumption |
| Subsistence | Extent of participation in subsistence-activities vs. wage labor | • Occupation or subsistence activities engaged in |
| Mobility (Orang Asli) | Access to the market economy | • Access to motorcycle or car<br>• Number of visits to Kuala Lumpur<br>• Number of visits to closest large town in last 2 weeks |
| Lifestyle categorization (Turkana) (adapted from Lea et al. 2020) | Delineation between Turkana living traditional, semi-traditional lifestyles, or highly market-integrated/urban lifestyles | • Turkana pastoralists defined as those solely practicing traditional pastoralism, relying on animal products, and eating traditional foods<br>• Peri-urban Turkana defined as those living in rural areas but not practicing traditional pastoralism<br>• Urban Turkana defined as those living in urban centers |

### Analysis of lifestyle and cardiometabolic health

We used linear and generalized linear models to analyze continuous and binomial cardiometabolic outcomes, respectively, performing modeling separately for each population. For each scale, we computed the amount of variance explained by comparing the difference in R^2^ between models with and without the focal scale included as a covariate. We then compared AIC between models with each scale for a given cardiometabolic outcome. For the three scales that best predicted cardiometabolic health in each population, we performed sensitivity analyses to determine the extent to which specific items within each scale contributed to variance in cardiometabolic health. In these sensitivity analyses, we removed one item in the scale and compared the difference in R^2^ between the model with the complete scale and the scale calculated with the item removed. We performed all analyses using the R computing language and RStudio version 4.3.2^47^. For all analyses, we performed a Benjamini-Hochberg correction for multiple hypothesis testing across all models (unless otherwise noted), using the *p.adjust* function in the R environment.

To understand whether lifestyle scales performed similarly among females and males, we repeated the modeling approach described above for these sexes individually. We then explicitly tested whether cardiometabolic phenotypes in females and males were differentially impacted by lifestyle variation by modeling sex by lifestyle interactions for each scale.

Next, we tested whether the items included in our nine scales could be decomposed into delineable axes of lifestyle variation. To do so, we used factor analysis, an unsupervised machine learning method, which is detailed in the supplement and was inspired by McDade and Adair 2001).

Lastly, we asked whether lifestyle variation, as quantified by each of our scales, showed monotonic associations with cardiometabolic health or whether these relationships might contain an inflection point at which the relationship between health and lifestyle changes. To test this, we modeled the relationship between each biomarker and each lifestyle scale with a linear and quadratic term, and determined the pairs for which a quadratic model was the better fit; for these pairs, we performed piecewise analyses to quantify inflection points in the relationship. These analyses are further described in the supplement.

## Results

### Changes in diet are distinct from changes in built environment during lifestyle transitions

We first generated eight metrics per population that encapsulated different facets of the lifestyle transition during urbanization and industrialization (Tables 1, S1; Figures 1B, S1). Using these scales, we quantified the transition from traditional, subsistence-level to industrialized, market-integrated lifestyles for both the Turkana and Orang Asli. Individuals included in the analysis span a wide lifestyle gradient in both countries (Figures 1C-E, S1).

We assessed the correlation between all scales within each population (Figure 1F-G). As the availability of highly processed foods generally occurs before larger-scale changes in infrastructure, occupation, education, and the accumulation of material wealth, we hypothesized that scales reflecting non-dietary facets of urbanicity would be more correlated to each other than to scales quantifying diet^24,31^. Indeed, scales capturing non-dietary lifestyle features such as the built environment, material wealth, and acculturation were more correlated to one another (mean Pearson r = 0.59 [Turkana], mean r = 0.58 [Orang Asli]) than to scales quantifying traditional or market-derived diet (mean r = 0.40 [Turkana], mean r = 0.28 [Orang Asli]) (t-test p = 1.71 x 10^-3^ [Turkana], t-test p = 5.53 x 10^-5^ [Orang Asli]; Figure 1F-G). Overall, lifestyle measures in Orang Asli trended towards being less correlated than in Turkana (t-test p = 0.24), suggesting that there may be more heterogeneity in how lifestyle is changing in Peninsular Malaysia than Turkana County and surrounding areas.

### Urbanicity predicts cardiometabolic health

Next, we sought to understand which features of lifestyle most strongly predicted our 16 cardiometabolic health phenotypes. As expected, we found that less traditional lifestyles (e.g., higher urbanicity, acculturation, integration to the market economy) were associated with larger waist circumference, higher weight, higher BMI, higher body fat percentage, higher triglycerides, and greater prevalence of obesity or overweight in both the Turkana and Orang Asli (Figure 2A-D; Tables S4-5). Among the Turkana, we also observed that more urban lifestyles were positively associated with higher blood pressure, higher LDL:HDL cholesterol, and higher glucose levels (Figure 2A; Table S4). As expected, we observed substantial heterogeneity in the amount of variance explained by lifestyle per cardiometabolic biomarker, with traits related to body composition being more highly explained by lifestyle than those related to blood pressure or lipids (Figure S2A-B).

**Figure 2.**
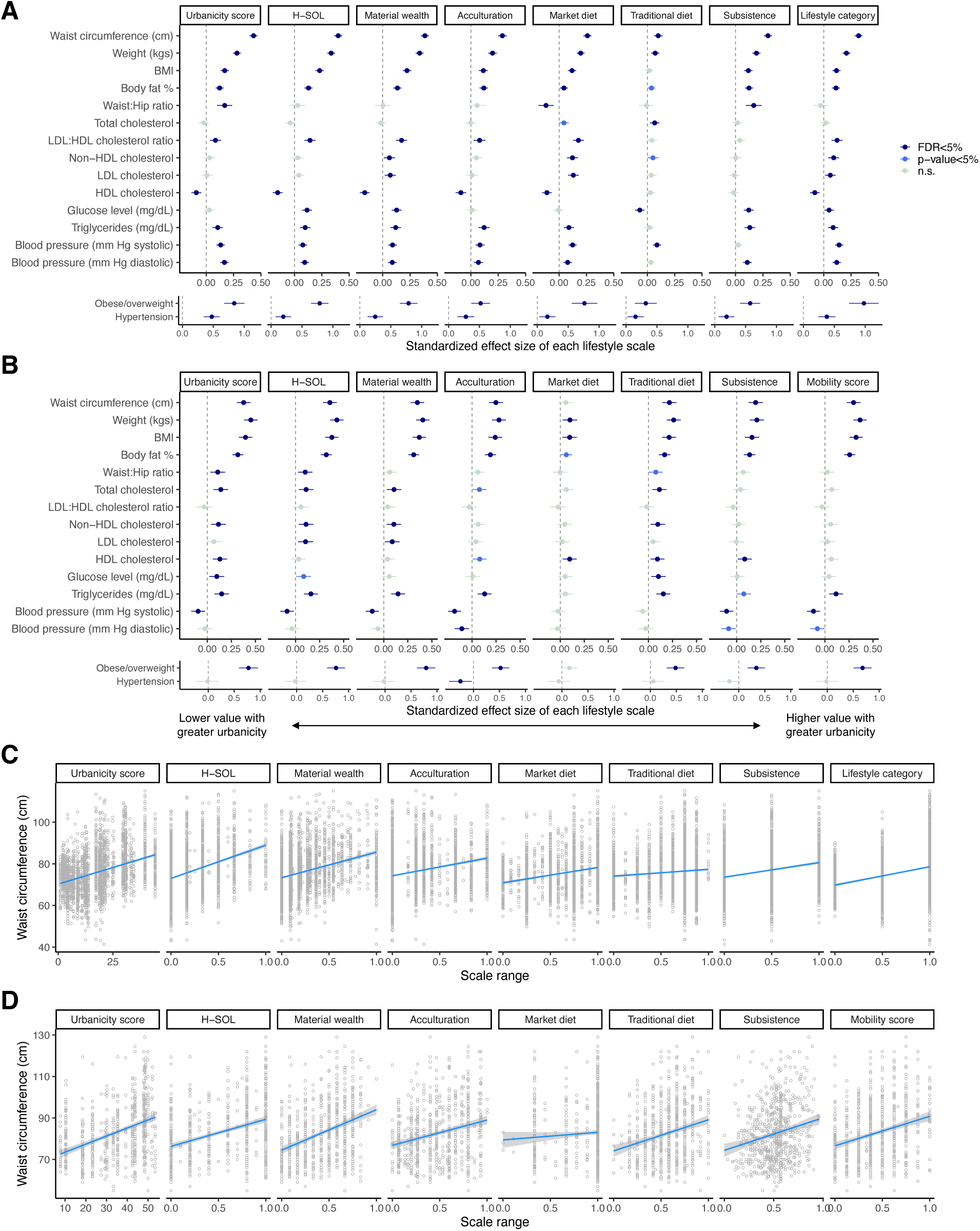
Effect of lifestyle on sixteen cardiometabolic health outcomes for both populations. Standardized effect sizes of lifestyle–as quantified by each of the nine scales–for all cardiometabolic traits in (A) Turkana and (B) Orang Asli. Dark blue points represent associations which passed an FDR of 5%, medium blue points represent associations with a significant p-value <0.05 but that did not pass a FDR of 5%, and light blue points represent non-significant associations. Waist circumference is positively associated with higher urbanicity as quantified by nearly all scales in (C) Turkana and (D) Orang Asli.

To determine which lifestyle scale explained the greatest amount of variance, we compared model fits for each cardiometabolic trait. We found that three lifestyle scales–all of which primarily measure material wealth and infrastructure–consistently best predicted cardiometabolic phenotypes and found strikingly high concordance between results for both populations (Figure 3A-B; Tables S4-5). When we assessed categories of cardiometabolic traits, we found that body composition phenotypes in Turkana and Orang Asli were best predicted by our location-based urbanicity score, material wealth scale, and H-SOL. Turkana blood biomarkers (i.e., lipids) were best predicted by our material wealth and market diet scales, while blood biomarkers in Orang Asli were best predicted by the location-based urbanicity score and the traditional diet scale. Blood pressure in Turkana was best predicted by location-based urbanicity score and lifestyle category, while blood pressure in Orang Asli was best predicted by our acculturation and subsistence scales (Figure 3A-B). Interestingly, we observed similarities in the scores that were generally least predictive in both populations, with these being diet-related scores and the subsistence score which measures the extent to which individuals participated in subsistence-level activities (e.g., pastoralism, foraging) in both populations (Figure 3A-B; Tables S4-5).

**Figure 3.**
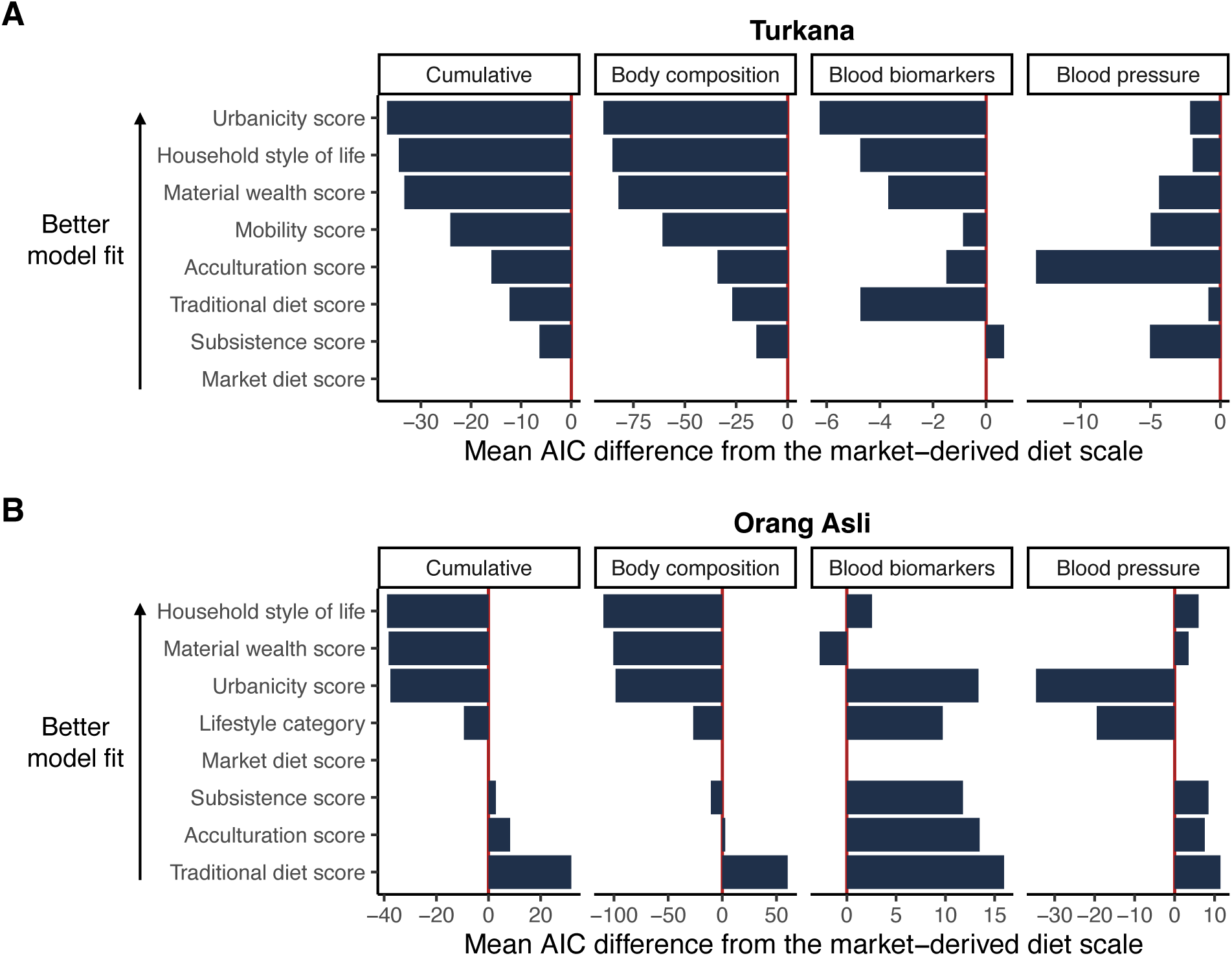
Scales that measure infrastructure and ‘Western’ material wealth better predict cardiometabolic biomarkers than scales that measure acculturation, subsistence/occupation, or diet. The mean difference in AIC from the market-derived diet scale across all cardiometabolic traits (cumulative), and traits associated with body composition, blood biomarkers, or blood pressure in (A) Turkana and (B) Orang Asli. The y-axis is ordered by lowest mean AIC across the four categories shown.

Further, we hypothesized that different extents of urbanicity, wealth, acculturation, or dietary changes might affect cardiometabolic health to varying degrees throughout the lifestyle transition. To test this, we investigated non-linearities in how lifestyle features are associated with cardiometabolic health in the Turkana and found evidence for many nonlinear relationships, adding nuance to these relationships that future studies should investigate (Figure S3A-B). Investigating these relationships can reveal, for example, that the slope of material wealth and waist circumference is positive at low levels of material wealth and negative at high levels of material wealth (Figure S3C). Understanding when in the lifestyle transition the association between lifestyle factors and cardiometabolic health shift may reveal when during the transition lifestyle interventions may be particularly impactful.

### Factor analyses consistently show that the built environment and diet act independently on cardiometabolic health across populations

We next asked whether the items included in our lifestyle scales decomposed onto delineable facets of lifestyle transition using an unsupervised machine learning method. To do so, we performed factor analyses for the Turkana and Orang Asli separately, using all constituent items in the eight urbanicity scales we quantified for each population. We determined that two factors accounted for the majority of the variance (factor 1 proportion of variance: 34% [Turkana], 43% [Orang Asli], factor 2 proportion of variance: 26% [Turkana], 19% [Orang Asli]) and were strikingly consistent between the two populations: the first factor was primarily associated with features related to infrastructure (e.g., electricity, plumbing) and the built environment (e.g., housing materials, material wealth) (Figures 4A, S4A-F). The second factor loaded most strongly onto features related to diet and was highly associated with the a priori market diet scale in both Turkana (r = 0.97) and Orang Asli (r = 0.98) (Figures 4A, S4B, S4D-F). We then modeled associations between each factor and the 16 cardiometabolic phenotypes and generally found expected relationships, for example that body composition measures were positively and additively associated with both factors for each population (Figure 4B; Tables S6-7). Together, these results recapitulated our findings from a priori defined lifestyle scales: features associated with the built environment are more predictive of cardiometabolic health than diet. This is notable because diet seemingly has more proximate effects on cardiometabolic health than structural environmental features. As such, these environmental factors likely serve as integrated and robust proxies for more proximate mechanisms by which urbanicity affects health.

**Figure 4.**
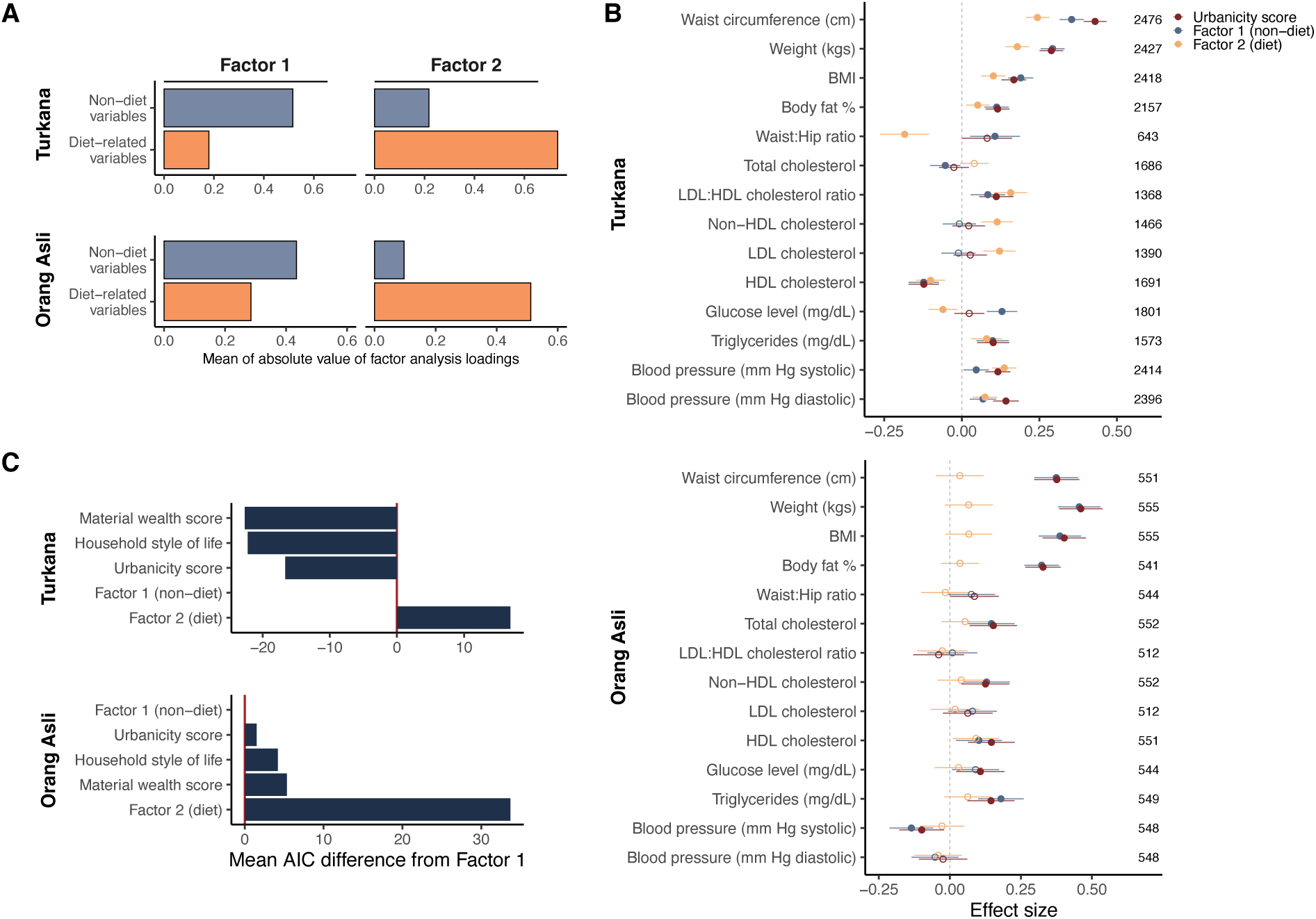
Factor analysis decomposes lifestyle into two axes: infrastructure and the built environment vs. diet. (A) Loadings for factors 1 and 2 in both populations capture highly similar features of the data, with factor 1 generally correlating with features of lifestyle not related to diet and factor 2 being most associated with diet-related variables. (B) Standardized effect sizes showing the effect of each factor (factors modeled separately) on each cardiometabolic trait. Effect sizes from models of the location-based urbanicity scale are shown for a comparison. Filled circles indicate statistically significant effects (FDR<5%) and open circles are not statistically significant. Sample sizes are shown on the right-hand side. (C) Mean difference in AIC across all continuous cardiometabolic traits for the top three lifestyle scales and the two factors from our factor analysis.

Next we sought to quantify whether the results of the factor analysis were more strongly predictive of cardiometabolic health than our a priori measures of lifestyle. To this end, we compared models with a covariate of factor 1 or factor 2 (modeled separately) to those with the top three performing lifestyle scales in each population (material wealth, H-SOL, and the urbanicity scale). In Turkana, the models of the three lifestyle scales had lower AIC scores across all health outcomes than either factor 1 or factor 2, with material wealth and H-SOL being the best performing measures and performing very similarly (Figure 4C; Table S6). For Orang Asli, factor 1 best predicted cardiometabolic health, with the urbanicity scale very closely being the second-best performing overall metric; these were followed by H-SOL, the material wealth score, and finally, factor 2 (Figure 4C; Table S7).

### Female cardiometabolic health is often more strongly impacted by urbanicity than male cardiometabolic health

We were interested in understanding whether (i) lifestyle variation affects cardiometabolic health in a sex-dependent manner and (ii) whether the measures of lifestyle that best predict cardiometabolic phenotypes are consistent between these sexes. We observed many sex by scale interactions, especially among Turkana where between two (market and traditional diet scales) to seven (acculturation scale) cardiometabolic phenotypes exhibited significant interaction effects, dependent on lifestyle scale (Figures 5A, S5A-B; Table S8). For example, focusing on the location-based urbanicity scale, we found that Turkana female cardiometabolic phenotypes were overwhelmingly more affected by urbanicity than males (binomial test: p = 4.18 x 10^-3^), especially for traits related to body composition (Figures 5A-B, S5A; Table S8). In Orang Asli, many cardiometabolic traits showed similar trends of sex by lifestyle effects but were not significant following multiple hypothesis testing correction (Figures 5A, S5B; Table S9). Additionally, we found that the rank-order of lifestyle scales was highly similar between the sexes in both populations, with the material wealth and community-level urbanicity scale best predicting cardiometabolic health in both sexes (Figure 5C; Tables S10-11). In both populations, scales measuring subsistence-participation and traditional diet were among the least predictive scales for cardiometabolic health in both sexes (Figure 5C; Tables S10-11).

**Figure 5.**
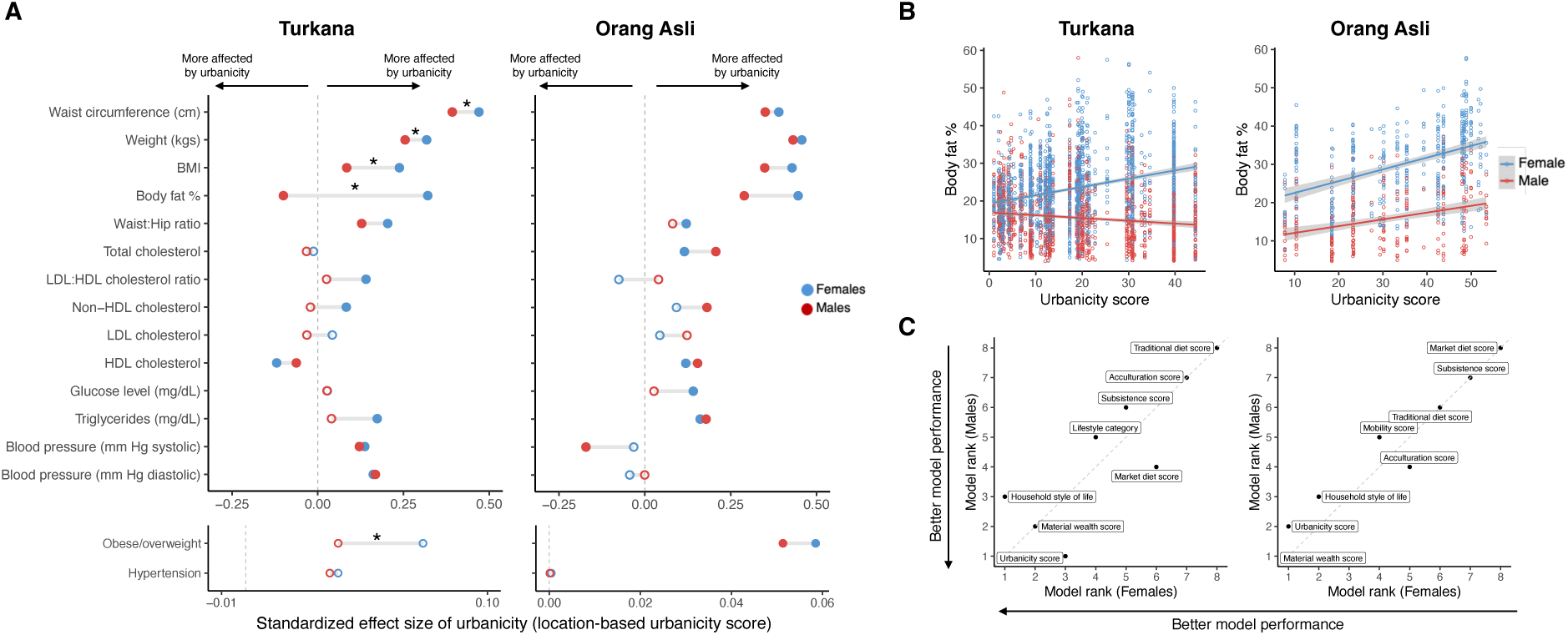
Female cardiometabolic health is generally more impacted by urbanicity than males. (A) Standardized effect size of urbanicity from models within females (blue) and males (red) for both populations. Points further from y=0 indicates a stronger effect of urbanicity. Stars denote significant sex x urbanicity interaction effects from the interaction models. Filled circles denote effects statistically significant at a FDR<5%, open circles are not significant at FDR<5%. (B) Example of interactive effects of sex and urbanicity (significant in Turkana, trending in Orang Asli). (C) Comparison of rank order of mean model fit between males and females across all cardiometabolic traits in both populations.

## Discussion

Indigenous peoples around the world have an average life expectancy approximately 10 years shorter than non-Indigenous individuals, largely due to non-communicable and cardiometabolic diseases, many of which increase with the transition from subsistence-level lifestyles to urban or market-integrated lifestyles^3,48^. Reducing health gaps for subsistence-level and Indigenous populations is a major goal of public health researchers, practitioners, and policy-makers worldwide, and the World Health Organization^3^. It is possible to conceive of many ways that changing lifestyles and environments influence cardiometabolic health of subsistence-level, Indigenous, or other transitioning populations. Possible mechanisms range from economic forces affecting physical activity, to altered dietary options, ethnic non-retention and acculturation, or features of the built environment and modern amenities that broadly shift behavioral patterns. Yet it remains unclear how these factors vary vis a vis in predicting downstream outcomes and few studies have attempted to directly compare different dimensions of lifestyle change in a way that could be feasibly applied in cross-cultural settings around the world. By integrating detailed anthropological and biomarker data from two transitioning subsistence-level, Indigenous societies, we demonstrate that lifestyle variation can be 1) meaningfully quantified in multiple dimensions that separate people living across wide gradients, 2) measured empirically such that cross-cultural comparison is feasible, and 3) linked directly with physiological data to determine how dimensions of lifestyle differentially impact health.

First, we found that despite lifestyle transitions manifesting very differently in Kenya and Peninsular Malaysia—the two populations we studied live under very different social, environmental, and political landscapes—lifestyle factors exhibit strikingly similar effects on cardiometabolic health in both populations. Together, these results show that quantitative measures of lifestyle are practically useful in distinct contexts, and may be generally useful for identifying vulnerable populations or designing intervention approaches, even across disparate environmental and socio-political contexts. Further, the structural and systemic reasons underpinning Indigenous peoples’ lifestyle change are diverse, from autonomous decisions to pursue wage labor or urban economic opportunities to forced resettlement and assimilation^3,49^. The lifestyle change experienced by Turkana and Orang Asli encompasses both of these reasons^9,18,40^, but nonetheless we again observe similar overall effects of lifestyle variation on health.

Second, we found that the built environment and material wealth more strongly predicted cardiometabolic health on average than measures of seemingly more proximate features, such as the extent to which individuals consume a traditional subsistence diet and/or a market-derived diet with greater sugar, salt, and oil (as seen also by Lea et al. [2020] in the Turkana, and Liebert et al. [2013] in the Shuar)^18,29^. As processed foods and market-derived additives are well established to have direct effects on blood lipid levels and adiposity^50,51^, we expect that our results reflect that individuals living in areas with urban infrastructure (e.g., roads, gridded electricity) have likely had access to market-integrated items for longer and/or in at higher quantities than individuals with less urban infrastructure. In support of this hypothesis, we find that Orang Asli exposed to higher urbanicity–who generally consume more processed foods– report significantly less change in the consumption of processed oil and sugar between adulthood and childhood (oil: β = −0.45, p = 9.90 x 10^-6^, sugar: β = −0.39, p = 1.87 x 10^-4^).

We also found that an unsupervised machine learning method decomposed lifestyle variation into two clear axes in both populations–the built environment and diet–recapitulating that these aspects of lifestyle change are commonly separable. In other words, lifestyle variation related to infrastructure and material wealth may often vary somewhat independently from features related to diet, and can therefore be separately targeted for monitoring or intervention. Further understanding the distinctions between these facets can illuminate how and when in the lifestyle transition interventions are likely to be most impactful. For example, in our data, waist circumference increases steadily across low to moderate levels of material wealth and then decreases at high levels of wealth, while waist circumference increases only slightly across much of the market diet transition but much more steeply at high levels of market diet consumption (Figure S3C-D).

Third, we find that female cardiometabolic health–especially body composition–was more affected by urbanization than that of males, recapitulating previous findings in Turkana and many populations, and highlighting an important intervention target of Indigenous health disparities^3,18,28^. One potential explanation is greater sex difference in activity levels with increasing urbanicity, with females reducing their average activity levels to a greater extent than males in many transitioning societies, especially in areas that become less walkable^21,52^. While females may be more impacted, reduced activity levels and increased time sedentary likely contribute to urban adiposity and cardiometabolic morbidity more generally in LMIC^22,52^. To fully address these gaps, in-depth anthropological research is needed to understand sex-differences in activity levels in traditional, subsistence-level societies and how sex-based social roles and activity levels change throughout the lifestyle transition.

Practically, our study offers several methods to quantify diverse facets of lifestyle at the individual- and community-levels. Public and Indigenous health professionals are increasingly working across populations and countries, and methods that perform robustly in different contexts will help inform health monitoring and intervention strategies. Further, the scales we test that captured infrastructure and material wealth performed better or comparably to the main factor of a factor analysis that included dozens of items and can be measured in 10 short survey questions (Table S12) and are thus time- and resource-efficient. As studies with Indigenous and subsistence-level communities are often in remote locations with limited resources, it may in some cases be beneficial to collect aggregate location-level data based on a representative sample of individuals rather than data for each individual. Capturing general environmental features of communities may also be less prone to respondent error or recall about individual-level behavior.

Our study has limitations. First, we acknowledge that we use several outcome measures that may have different relationships to morbidity or mortality in our participant populations relative to the high income contexts in which they have been validated and in which they are commonly used, namely continuous BMI, obesity (yes/no), and hypertension (yes/no)^53^. Second, we acknowledge that there is mixed evidence that BMI under 30 (i.e., the threshold for obesity categorization) negatively impacts health^54,55^. As BMI is systematically biased by height^56^, comparisons of BMI between populations with substantially different population-average heights is not ideal. However, within-population BMI comparisons can be useful as a coarse measure of body composition and we include BMI in our analyses because it has been used in many previous studies and allows our results to be compared to prior work. Third, we note that while many of the lifestyle scales we present here have the same range, these scales are relative representations of the lifestyle transition and values will change depending on the items included in each scale. Scales may need to be adjusted to reflect traditional practices which differ between populations, for example quantifying pastoralism vs. farming and foraging in the two populations we present here. The scales we present are not meant to quantify equivalent positions in the lifestyle transition, but instead are useful in contexts in which there is variation across the lifestyle gradient and/or between populations. Finally, while studying two unrelated populations that live in very different environments provides strong evidence for the generalizability of our findings, repeating this investigation in additional transitioning populations will further illuminate the factors that drive context-dependency of lifestyle effects on cardiometabolic health.

In conclusion, we find a diverse set of 16 cardiometabolic phenotypes were similarly affected by lifestyle features in two very different Indigenous populations spanning extreme lifestyle gradients, and that infrastructure and material wealth exerted stronger effects on cardiometabolic health than diet in both populations. We also find that these results are consistent across females and males, though female body composition was generally more affected than males. While we did not formally investigate additional proximate factors that might moderate the relationship between lifestyle change and cardiometabolic health, the extensive variation we observe in health among individuals with similar lifestyles suggests that many factors join to determine inter-individual variation. Future work could identify the individual-level factors that predict vulnerability versus resilience during lifestyle transitions, for example physical activity levels^21,52^, sleep quality^57^, early life experiences^58^, and social connectedness and other sources of social protection^59^. Additionally, our results show that lifestyle does not always impact cardiometabolic health in a linear manner, and future work should identify the factors that modify the slopes of the relationship between lifestyle and cardiometabolic health. In summary, our work provides public health researchers a unified framework for quantifying different facets of the lifestyle transition that is comparable across populations, and our findings highlight that lifestyle change–especially increased urbanization– has extensive effects on cardiometabolic health across populations, contexts, sexes, and is a major target for intervention.

### Data sharing

Our highest priority, and that of the Turkana Health and Genomics Project and the Orang Asli Health and Lifeways Project, is to minimize risk to study participants. As such, both projects adhere to the “CARE Principles for Indigenous Data Governance” (Collective Benefit, Authority to Control, Responsibility, and Ethics) and are committed to the “FAIR Guiding Principles for scientific data management and stewardship” (Findable, Accessible, Interoperable, Reusable). To adhere to these principles while minimizing risks, individual-level data are stored in the protected data repositories and are available through restricted access. Requests for de-identified, individual-level data should take the form of an application that details the exact uses of the data and the research questions to be addressed, procedures that will be employed for data security and individual privacy, potential benefits to the study communities, and procedures for assessing and minimizing stigmatizing interpretations of the research results. Requests for de-identified, individual-level data will require institutional IRB approval (even if exempt). THGP and OA HeLP are committed to open science and the project leadership is available to assist interested investigators in preparing data access requests (see turkanahgp.com and orangaslihealth.org for further details and contact information). Data needed to evaluate the conclusions in the paper are present in the paper and/or the Supplementary Materials. Code used to generate lifestyle scales and to perform analyses described herein is available on our GitHub (https://github.com/mwatowich/Multi-population_lifestyle_scales).

## Data Availability

All data produced in the present study are available upon reasonable request to the authors

https://github.com/mwatowich/Multi-population_lifestyle_scales

## Acknowledgments

First and foremost, we thank Turkana and Orang Asli community members and leaders for their contributions and support of our scientific work. We also thank all previous members of the Turkana Health and Genomics Project and the Orang Asli Health and Lifeways Project. We are also grateful to the staff of Mpala Research Centre for their essential support.

## Supplementary Tables

1. Scale construction
2. Turkana demographic summary
3. Orang Asli demographic summary
4. Model results of health as a function of lifestyle scale (Turkana)
5. Model results of health as a function of lifestyle scale (Orang Asli)
6. Model results of cardio health as a function of factor 1 or 2 (Turkana)
7. Model results of cardio health as a function of factor 1 or 2 (Orang Asli)
8. Sex*lifestyle models (Turkana)
9. Sex*lifestyle models (Orang Asli)
10. Sex-specific lifestyle models (Turkana)
11. Sex-specific lifestyle models (Orang Asli)
12. Ten survey questions to generate infrastructure and material wealth scales

