## Supplemental Text and Figures for "The built environment is more predictive of cardiometabolic health than other aspects of lifestyle in two rapidly transitioning Indigenous populations"

The supplement consists of:

Supplementary Methods

Supplementary Results

Supplementary Figures 1-6

Supplementary References

### Supplementary Methods

#### Sampling overview

Data from Turkana were collected between March 2018 and November 2023 throughout Kenya as part of the Turkana Health and Genomics Project (THGP)[1]. Researchers traveled to locations where individuals of Turkana ancestry were known to reside and invited healthy adults (>18 years old) to take part in the study. Following approval from Turkana leaders and a community-wide discussion about the project goals, individuals who wished to participate had a structured interview with a research team member who was familiar with the local community and language of the participant (e.g., Turkana, Swahili, or English) and had basic anthropometric and health measurements taken (e.g., height, weight, blood pressure). Written, informed consent was obtained from all participants after the study goals, sampling procedures, and potential risks were explained to participants in their native language by researchers. Sampling procedures of the THGP are detailed further in Lea et al. (2020). The THGP was approved by the Princeton University Institutional Review Board for Human Subjects Research (Institutional Review Board no. 10237) and Maseno University (approval number MUERC-00519-18).

Data were collected from Orang Asli individuals in communities throughout Peninsular Malaysia between March 2020 and March 2024 as part of the Orang Asli Health and Lifeways Project (OA HeLP)[2]. Researchers traveled to communities which spanned a gradient of rural, remote communities to acculturated, market-integrated urban areas. Sampling locations were selected to include individuals from the three major Orang Asli ethnolinguistic groups. In each location, healthy adults (≥18 years old) were invited to participate in the study as part of the OA HeLP[2]. At each sampling location, permission to conduct research was obtained from community leaders and public forums were held to discuss the research, methods and procedures, potential risks, and participant compensation. Researchers explained the study goals, sampling procedures, and potential risks to participants in their native language and written, informed consent was obtained by all participants. Consenting adults participated in structured interviews with a research team member who was familiar with the local community and language of the participant (e.g., Malay), measurement of anthropometric phenotypes, and were invited to provide a blood sample. The OA HeLP was approved by the Medical Review and Ethics Committee of the Malaysian Ministry of Health (protocol ID: NMRR-20-2214-55565), the Malaysian Department of Orang Asli Development (permit ID: JAKOA.PP.30.052 JLD 21 (98)) and the Institutional Review Board of Vanderbilt University (protocol ID: 212175).

#### Cardiometabolic biomarker data collection

We measured 16 cardiometabolic phenotypes in the Turkana and Orang Asli, including anthropometric measurements, blood lipids, and blood pressure. Body composition measures included waist circumference, waist-to-hip circumference ratio, body fat percentage, weight, BMI, and overweight or obese (BMI ≥ 25) categorization. Anthropometric measures of body composition were measured using standard techniques detailed in Lea et al. (2020) and Wallace et al. (2022)[1,2]. For Turkana, waist and hip circumference measurements were generated by taking the mean of three measurements of each phenotype. Body fat percentage was measured using the Omron HBF-306C Handheld Body Fat Loss Monitor (Kenya) and a digital bioelectrical impedance scale (TANITA BC-558 FDA Cleared Ironman Segmental Body Composition Monitor) (Malaysia)[1,2]. Systolic and diastolic blood pressure were measured using the Omron 10 Series Wireless Upper Arm Blood Pressure Monitor (Kenya) and the Omron 5 Series Upper Arm Blood Pressure Monitor (Malaysia) following standard procedures. We categorized hypertension risk if systolic blood pressure was greater than 135 and diastolic blood pressure was greater than 85. Blood pressure was measured twice to reduce the effects of 'white coat hypertension' and we used the second measurement in our analyses.

In both studies, participants were invited to provide a small (approximately 15 mL) venous blood sample which we used to measure total cholesterol, triglycerides, and high- and low-density lipoproteins (collection further detailed in Lea et al. (2020). Lipids and blood glucose were measured from a 1mL venous blood sample using the Cardio Check Plus Analyzer panel from PTS Diagnostics[2]. LDL was calculated using the Friedewald Equation (LDL cholesterol = total cholesterol - HDL cholesterol – triglycerides/5) and non-HDL cholesterol was measured by subtracting HDL from total cholesterol[1]. We removed any cholesterol values under 50 mg/dL from analysis as the Cardio Check Plus panel cannot reliably detect triglycerides or total cholesterol under this threshold.

### Data filtering

We removed individuals from analyses if (i) they were missing data for age or sex, (ii) they were pregnant, or (iii) they were taking medications for hypertension or diabetes in Turkana for which we had these data (n = 4). For all numeric cardiometabolic traits, we removed outliers greater than 5 standard deviations away from the mean as these likely represent measurement error or individuals with unusual physiology. Before statistical analyses, all predictor variables and continuous biomarkers were mean centered and scaled by their standard deviation (SD), using the "scale" function in R. Consequently, all reported effect sizes are standardized and represent the effect of a given variable on the outcome in terms of increases in SDs. Participant sex was characterized as that assigned at birth and we analyzed sex as a binary variable (i.e., female/male). We acknowledge that sex consists of a suite of biological attributes and that binary assignment and analysis may not reflect the full diversity of this complexity within our participant sample.

### Development of a priori lifestyle domain scales

We developed and quantified six scales based on a priori knowledge of different facets of the lifestyle transition. These included: material wealth, subsistence strategy, acculturation, traditional diet, market-derived diet, and mobility.

In highly industrialized societies, higher material wealth is associated with better cardiometabolic health (e.g., hypothesized stage five of the nutritional transition), but in LMIC contexts, higher wealth often correlates with higher risk of diabetes and adiposity[3]. We quantify material wealth specifically in the context of 'Western' or industrialized concept of wealth that measures market-based assets accumulated through the cash economy, including the quantification of ownership of or access to a mobile phone, monetarily expensive household building materials, and larger household size. In both populations with which we work, we expect that these items measure participation in the cash economy and general economic capital[4].

The subsistence strategy domain aims to capture the extent to which individuals participate in traditional subsistence agriculture or hunting or gathering, or wage-labor. As many individuals use multiple strategies, we measure subsistence strategy in a continuous manner that quantifies effort in the market economy on a scale from no participation to full participation.

Within LMIC, cardiometabolic health is generally negatively associated with educational attainment, likely because this correlates with urban living and socioeconomic status[3,5–7]. Educational attainment is a primary conduit to acculturation and assimilation with majority cultures, which is also indicated via proficiency in the majority or national language and knowledge of majority customs[8].

Changes in diet often co-occur with lifestyle changes in acculturation, occupation, wealth, and the built environment during urbanization, making it difficult to disentangle their specific effects[6]. Specifically, dietary transitions include reduced consumption of or reliance on foods traditionally eaten by the subsistence-level population and an increased consumption of foods derived from the cash economy or global market[9–11]. Market-derived foods are often higher in processed carbohydrates, sugars, and fats than traditional foods which tend to be plant-based or unprocessed meats. Market-derived foods have been extensively connected to poor cardiometabolic health outcomes[6,12].

Additionally, for Orang Asli only, we developed a scale of individuals' mobility and frequency of travel to urban areas. We develop the mobility scale for Orang Asli and not for Turkana because we expected that vehicle access in Malaysia is more indicative of market access because driving to reach areas with greater infrastructure is often feasible within a single day, while in Kenya, urban centers are a multi-day drive from traditional Turkana lands.

### Factor analysis

During the transition from traditional, subsistence-level lifestyles to market-integration and industrialization, many facets of the environment change simultaneously, meaning that many of the features of lifestyle we measured are inherently correlated. We therefore performed factor analyses in both populations to characterize (i) axes of variation that explain cardiometabolic health in each population and (ii) quantify the environmental features that most strongly predict cardiometabolic health in both populations. To do so, we included all of the features of urbanization, acculturation, diet, and market-integration that were used in the generation of any of our eight individuals-level lifestyle scales (i.e., not community-level) in a factor analysis. We removed three features for Turkana (languages spoken, blood consumption, and fermented milk consumption) and one feature (religion practiced) for Orang Asli that had more than double the missingness of any other feature. We performed the factor analysis using the *fa* function in the R psych package with the varimax rotation to produce orthogonal factors[13]. Scree plots revealed that two factors explained the majority of the variance among lifestyle features in the Turkana and Orang Asli (Figure S4A-B). After quantifying two factors in each population, we investigated the loadings of each feature included in the factor analysis, and modeled the effect of each resulting factor on all cardiometabolic outcomes (Figure S3C-D). To compare whether the factor analysis or a priori urbanicity scales better predicted cardiometabolic phenotypes, we compared the AIC of models with each factor (factor one or factor two) to the models of the top three urbanicity scales, for each population.

### Piecewise analyses

We next investigated whether the relationship between cardiometabolic health and lifestyle was consistent across the lifestyle gradient we observed. Specifically, we (i) assessed whether the relationship between cardiometabolic health and lifestyle was better described by linear or quadratic functions of lifestyle and (ii) quantified inflections in the relationship between urbanicity and cardiometabolic health. We explored these relationships only in Turkana, as our Orang Asli sample size limited detection of non-linear effects. First, using the same modeling schema as described above, we modeled six lifestyle scales as a linear and quadratic term. We did not model the Turkana lifestyle categorization scale from Lea et al. (2020) or the subsistence scale because these each had low variation with individuals being classified into three categories, resulting in non-convergence of downstream piecewise analyses. Following modeling of all remaining lifestyle scales and biomarkers, we compared model fit (AIC<2) between models with the quadratic term of lifestyle to those with only the linear term and which the non-linear effect of lifestyle was significant. Forty-four models passed these criteria. After determining biomarker/lifestyle scale pairs for which the quadratic term fit better, we performed piecewise analysis using the R package segmented to quantify the different relationships present for these pairings and the inflection point at which the slopes change[14]. As the piecewise analysis via the segmented package requires a starting value estimate of inflection, we transformed the urbanicity scale to range from 0 to 1 (all other scales already had this range), and set the psi to 0.5 for all models. We controlled for age and sex in all piecewise models. Finally, we used linear models to test whether inflection points differed depending on biomarker type (e.g., blood pressure, body composition) or lifestyle scale.

### Supplementary Results

#### Sensitivity analyses reveal consistency in urbanicity features that best predict cardiometabolic health

We observed striking consistency in the scales that best predicted cardiometabolic health between Turkana and Orang Asli and we therefore sought to uncover the items within each scale that most explained these outcomes. To this end, we performed sensitivity analyses for each of these three measures–our material wealth index, H-SOL, and location-based urbanicity scale–by removing one item in the scale and calculating the difference in $R^2$ between the reduced-item scale and the complete scale (Figure S6A-C). As expected, our sensitivity analyses revealed that removing items from these three scores generally resulted in slightly lower absolute $r^2$ in the reduced model (Turkana: range $R^2$ difference = -0.01-0.02, mean $R^2$ difference = $9.27 \times 10^{-5}$, Orang Asli: range $R^2$ difference = -0.01-0.06, mean $R^2$ difference = $2.36 \times 10^{-4}$; Figure S6A, S6C). We did not observe that the removal of any particular item from any of the three scales we tested resulted in consistently better model fit (Figure S6A, S6C). We expect that these results reflect the reality that the items that comprise these three scores are highly correlated and likely capturing very similar variation in lifestyle, thus the removal of any one item will not severely alter the lifestyle variation that the score can detect.

#### Non-linear relationships between lifestyle scales and cardiometabolic biomarkers

To quantify whether cardiometabolic traits varied linearly or nonlinearly across the lifestyle gradient we observed in Turkana, we compared models with lifestyle scales modeled as a linear or quadratic variable (Table S13). We found forty-four lifestyle scale/biomarker pairs that showed non-linear relationships and quantified inflection points at which the relationship between lifestyle and cardiometabolic health changed. We observed substantial variation in the point at which inflections occurred along the lifestyle gradient (Figure S3A). First, we tested whether different biomarker types (e.g., body composition, blood pressure, blood lipids/biomarkers) differed in the proportion of the lifestyle gradient at which inflection points occurred. We found no significant difference in inflection estimate by biomarker group. We then asked whether different lifestyle scales were associated with earlier or later inflections of cardiometabolic health. We found that the location-based urbanicity score was associated with earlier inflections in cardiometabolic health than traditional diet and market-derived diet (p-values of linear model with urbanicity score as the reference group vs: market diet = 0.02, traditional diet = 0.01; Figure S3B; Table S14). Investigating individual relationships between lifestyle scales and cardiometabolic biomarkers can reveal nuances that linear relationships or aggregate analyses can overlook, for example, the relationships we describe in Results section "Urbanicity predicts cardiometabolic health" which are shown in Figures S3C-D.

### Supplementary Figures

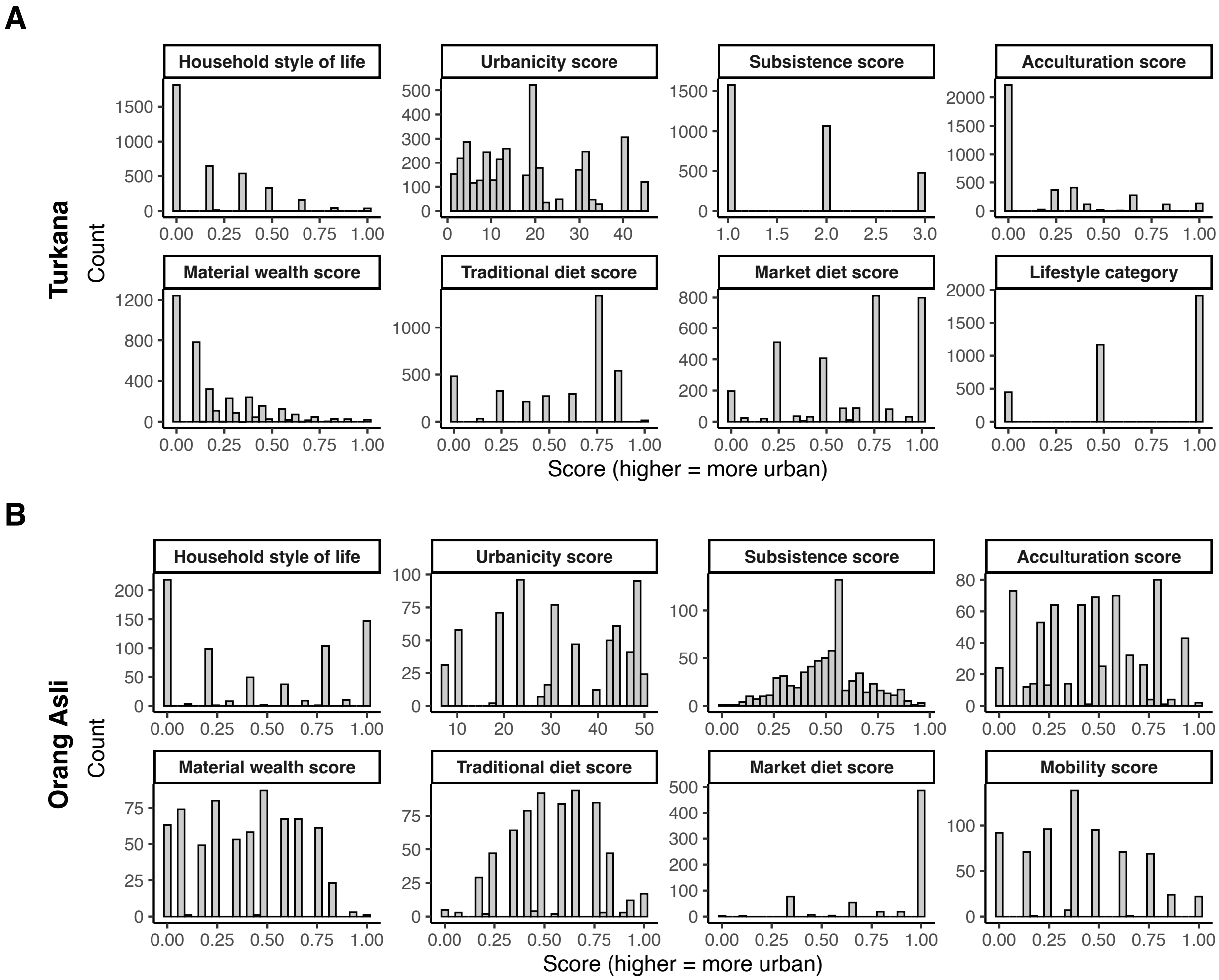


**Figure S1:** Histograms of all lifestyle scale scores for (A) Turkana and (B) Orang Asli.

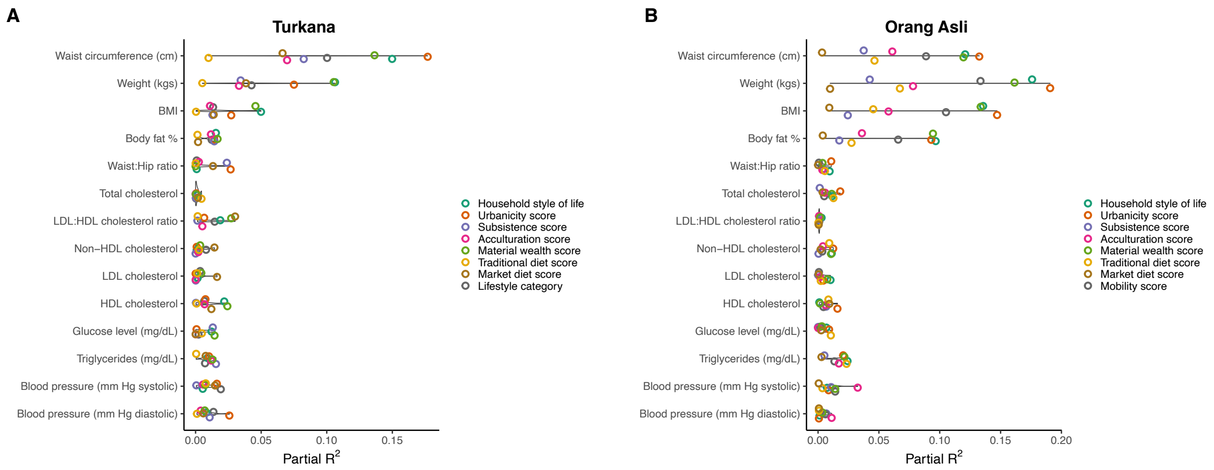


**Figure S2:** $R^2$ from models of each cardiometabolic biomarker and lifestyle scale for (A) Turkana and (B) Orang Asli.

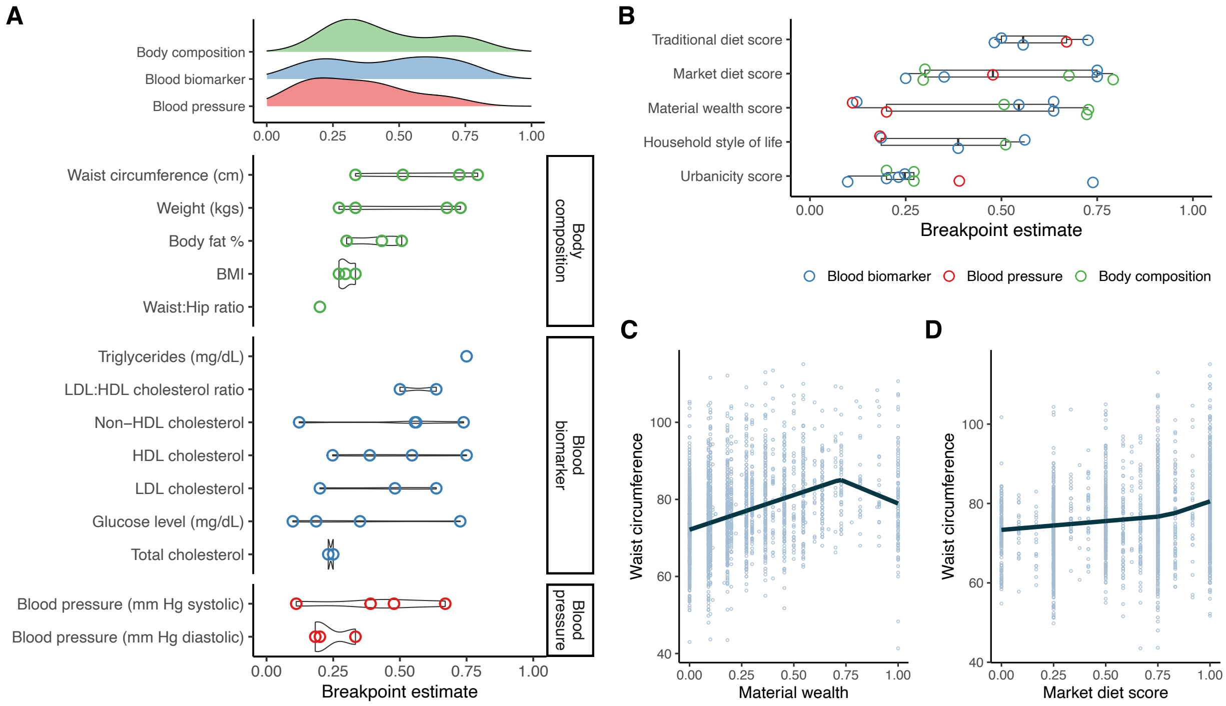


**Figure S3:** Piecewise analyses reveal inflection points in how lifestyle variation is associated with cardiometabolic health in Turkana. (A) Estimates of inflection points from piecewise analyses of different cardiometabolic biomarkers. (B) Estimates of inflection points from piecewise analyses of different cardiometabolic biomarkers by lifestyle scale. (C) Example of bi-slope relationship between material wealth scale and waist circumference, line plots fitted values from the piecewise model. (D) Example relationship between the market-derived diet scale and waist circumference.

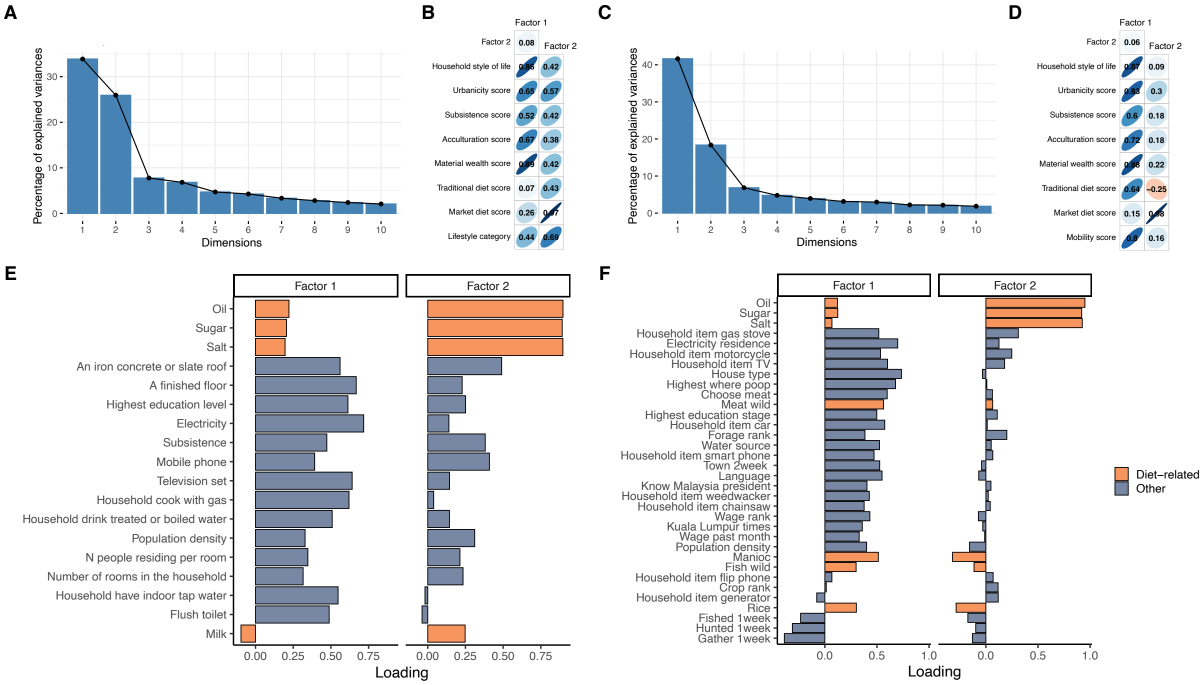

**Figure S4:** Factor analyses reveal that lifestyle variation decomposes into non-diet and diet-related axes. Scree plots from all items in (A) Turkana factor analysis and (B) correlation of factors 1 and 2 with all Turkana lifestyle scales. Scree plots from all items in (C) Orang Asli factor analysis and (D) correlation of factors 1 and 2 with all Orang Asli lifestyle scales. Loading scores for all items included in the factor analyses for (E) Turkana and (F) Orang Asli.

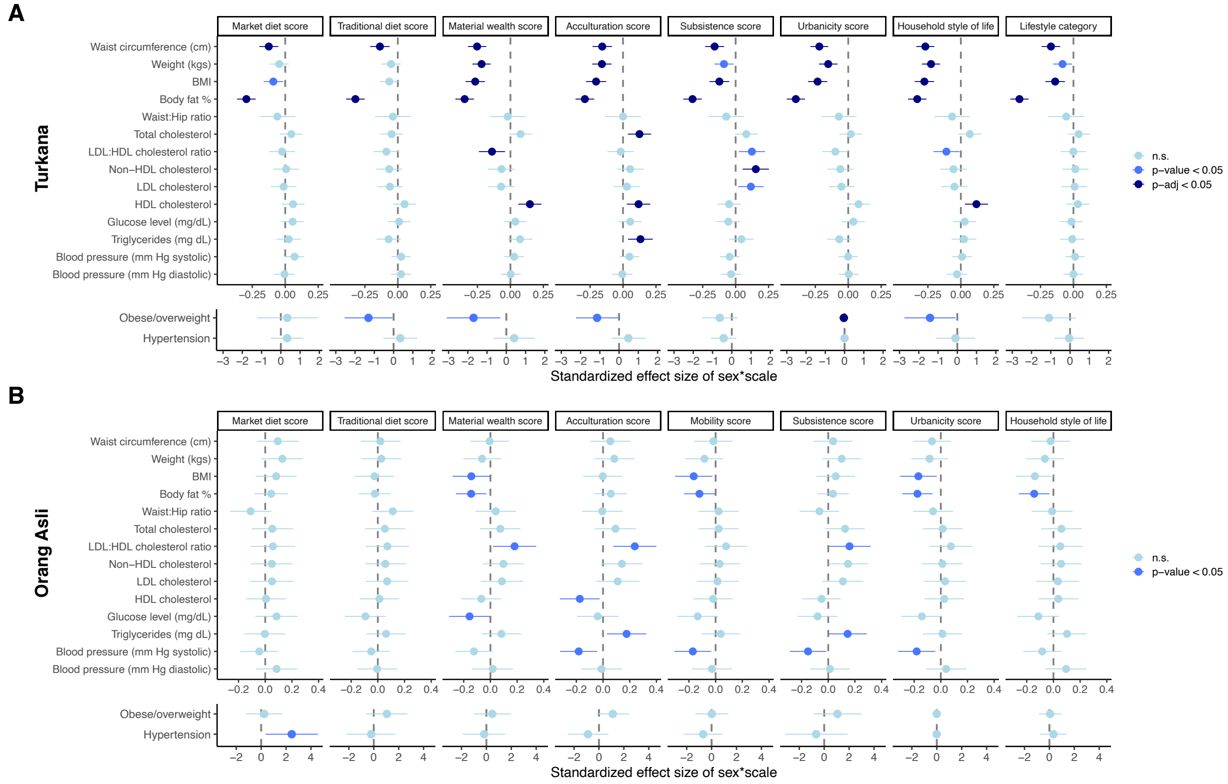


**Figure S5:** Standardized effect sizes of sex by lifestyle scale for (A) Turkana and (B) Orang Asli. Sex is categorized as female=0 and male=1, such that more negative values generally indicate greater sex by lifestyle interactions in females. One notable exception is HDL, which is generally negatively associated with lifestyle change (especially in Turkana), such that positive effect sizes indicate female HDL is more strongly negatively associated with urbanicity than males.

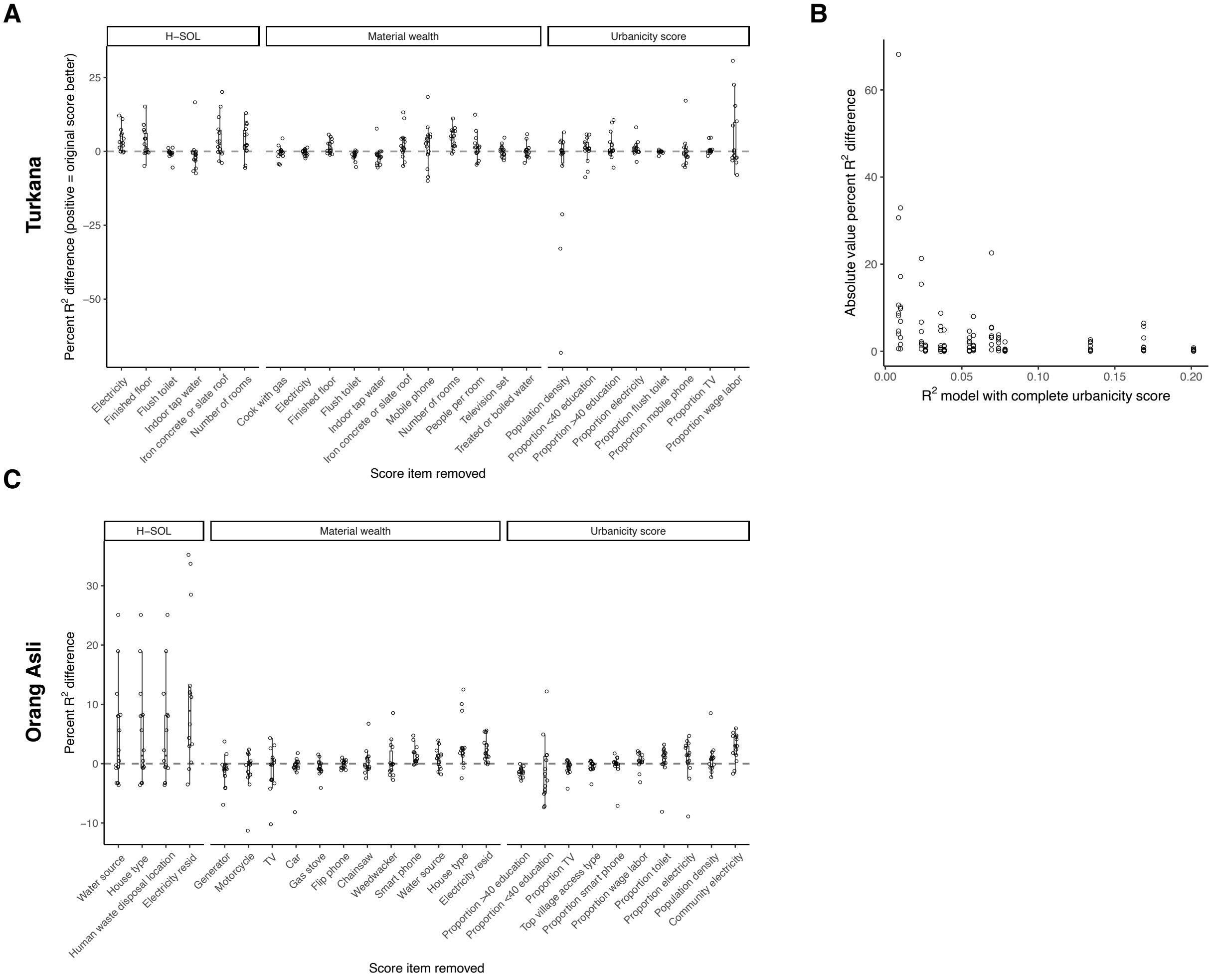


**Figure S6:** Sensitivity analyses show consistency in score performance. Sensitivity analyses iteratively removed one item from each of the top three lifestyle scores and assessed performance via the percent change in model fit (percent change $R^2$). (A) Percent difference in partial $R^2$ between models for Turkana. (B) Plot of $R^2$ from the model with the complete location-based urbanicity score (x-axis) and the absolute value of the percent difference of $R^2$. (C) Percent difference in partial $R^2$ between models for Orang Asli.
